# Using random testing in a feedback-control loop to manage a safe exit from the COVID-19 lockdown

**DOI:** 10.1101/2020.04.09.20059360

**Authors:** Markus Müller, Peter M. Derlet, Christopher Mudry, Gabriel Aeppli

## Abstract

We argue that frequent sampling of the fraction of infected people (either by random testing or by analysis of sewage water), is central to managing the COVID-19 pandemic because it both measures in real time the key variable controlled by restrictive measures, and anticipates the load on the healthcare system due to progression of the disease. Knowledge of random testing outcomes will (i) significantly improve the predictability of the pandemic, (ii) allow informed and optimized decisions on how to modify restrictive measures, with much shorter delay times than the present ones, and (iii) enable the real-time assessment of the efficiency of new means to reduce transmission rates.

Here we suggest, irrespective of the size of a suitably homogeneous population, a conservative estimate of 15’000 for the number of randomly tested people per day which will suffice to obtain reliable data about the current fraction of infections and its evolution in time, thus enabling close to real-time assessment of the quantitative effect of restrictive measures. Still higher testing capacity permits detection of geographical differences in spreading rates. Furthermore and most importantly, with daily sampling in place, a reboot could be attempted while the fraction of infected people is still an order of magnitude higher than the level required for a relaxation of restrictions with testing focused on symptomatic individuals. This is demonstrated by considering a feedback and control model of mitigation where the feed-back is derived from noisy sampling data.

## I. INTRODUCTION

The COVID-19 pandemic has led to a worldwide shut-down of a major part of our economic and social activities. This political measure was strongly suggested by epidemiologic studies assessing the cost in human lives depending on different possible policies (doing nothing, mitigation, suppression) [1–4]. Mitigation can be achieved by combinations of different measures, including physical distancing, contact tracing, restricting public gatherings, and the closing of schools, but also the testing for infections. The quantitative impact of very frequent testing of the entire population for infectiousness has been studied in a recent unpublished work by Jenny et al. in Ref. [5]. We will estimate in Sec. III that to fully suppress the COVID-19 pandemic by widespread testing for infections, one needs a capacity to test millions of people per day in Switzerland. This should be compared to the present number of 7’000 tests per day across Switzerland. ^1^ Here we suggest that by testing a much smaller number of randomly selected people per day one can obtain important quantitative information on the rates of transmission, so as to enable well-informed decisions.

Figure 1 summarizes the key concept of the paper, namely a feedback and control model for the pandemic. The essential output from random testing is the growth rate of the number of currently infected people, which it-self is regulated by measures such as those enforcing physical distances between persons (physical distancing), ^2^ and whose tolerable values are fixed by the capacity of the health-care system. A feedback and control approach[6], familiar from everyday implementations such as thermostats regulating heaters and air conditioners, should allow policy makers to damp out oscillations in disease incidence which could lead to peaks in stress on the healthcare system as well as the wider economy. Any other measurement of the fraction of currently infected people can replace the random testing, for example there are proposals to estimate this fraction from analysis of sewage water with PCR tests [7, 8].

**FIG. 1.**
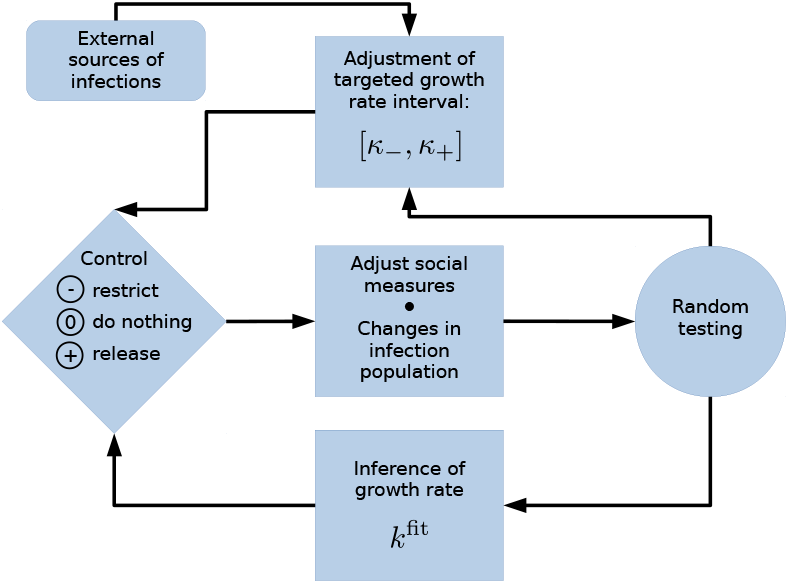
Feedback and control loop that allows stabilization of the pandemic. The key quantity measured by random testing is the growth rate *k* of infection numbers. If *k* exceeds a tolerable upper threshold *κ*_+_, restrictions are imposed. For *k* below a lower threshold *κ*_−_, and if infection numbers are below critical, restrictions are released. In the absence of a substantial influx of infected people from outside the country, and provided infection numbers are below a critical value, the optimal target of the growth rate is *k* = 0, corresponding to a marginally stable state, where infections neither grow nor decrease exponentially with time. If higher testing rates are available, the measured observables and control strategies can be geographically refined, particularly to avoid hotspots.

An important further benefit of our feedback and control scheme is that it allows a much faster and safer reboot of the economy than with feedback through confirmed infection numbers of symptomatic persons or deaths [4], for the latter, secondary indicator is heavily delayed and reflects the state of the pandemic only incompletely as it does not account for asymptomatic carriers. Figure 2 illustrates the resulting difference in the ability to control the disease.

**FIG. 2.**
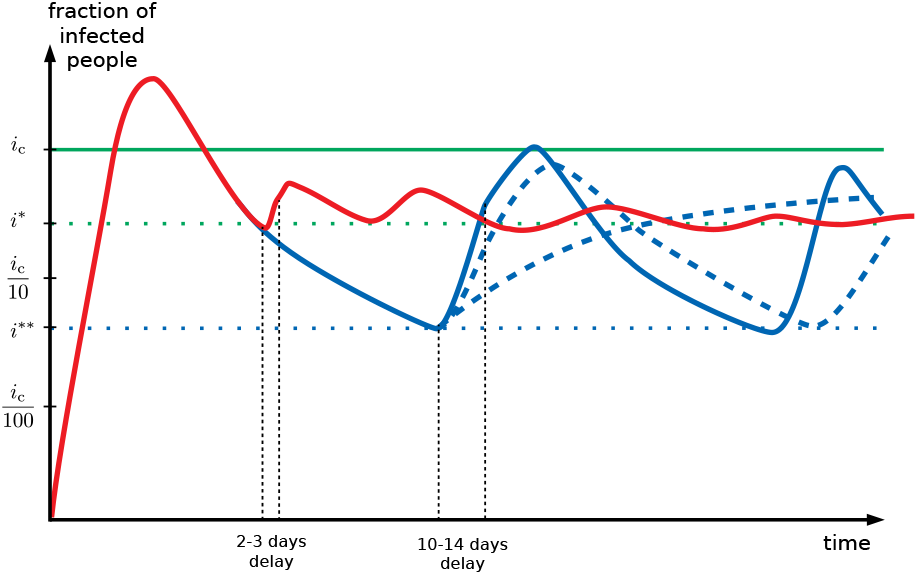
Dynamics of the pandemic with and without a feedback and control scheme in place, as measured by the fraction *i* of currently infected people (logarithmic scale). After the limit of the health system, *i*_c_, has been reached, a lockdown brings *i* down again. The exponential rate of decrease is expected to be very slow, unless extreme measures are imposed. The release of measures upon a reboot is likely to re-induce exponential growth, but with a rate difficult to predict. Three possible outcomes are shown in blue curves in the scenario without testing feedback, where the effect of the new measures becomes visible only after a delay of 10-14 days. In the worst case, *i* grows by a multiplicative factor of order 20 before the growth is detected. A reboot can thus be risked only once *i* ≤ *i * \**≡ *i*_c_*/*20, implying a very long time in lockdown after the initial peak. Due to the long delay until policy changes show observable effects, the fluctuations of *i* will be large. Random testing (the red curve) has a major advantage. It measures *i* instantaneously and detects its growth rate within few days, whereby the higher the testing rate the faster the detection. Policy adjustments can thus be made much faster, with smaller oscillations of *i*. A safe reboot is then possible much earlier, at the level of *i* ≤ *i*^*\**^ ≈ *i*_c_*/*4.

Without feedback and control informed by a primary indicator, analogous to the temperature provided by the thermometer in the thermostat example, measurable in (near) real time, there is a huge lapse between policy changes and the observable changes in numbers of positively tested people. To relax restrictions safely, the fraction of currently infected people must decrease to a level *i*^*\*\**^ such that a subsequent undetected growth during 10-14 days will not move it above the critical fraction *i*_c_ manageable by the health-care system. The current situation where we are mainly looking at lagging secondary indicators, namely infection rates among symptomatic individuals or even deaths, is comparable to driving a car from the back seat and with knowledge only of the damage caused by previous collisions. To minimize harm to the occupants of the vehicle, driving very slowly is essential, and oscillations from a straight course are likely to be large.

Daily random testing reduces the delay between changes in policy and the observation of their effects very significantly. Moreover, it directly measures the key quantity of interest, namely the fraction of currently infected people and its growth rate, information that is very valuable to gauge further interventions. Such information is much harder to infer from data about positively tested patients only, by fitting it to specific epidemiological models with their inherent uncertainties. The shortened time delay due to feedback and control allows a reboot to be attempted at much higher levels of infections, *i*^*\**^ > *i*^*\*\**^, which implies a much shorter time in lockdown.

We point out before proceeding further that this is a contribution from physicists that makes simplifying assumptions inconsistent with details of medical and epidemiological reality to obtain some key estimates and illustrate the basic principles of feedback and control as applied to the current pandemics. When reduced to practice, special attention will need to be paid to all aspects of the testing methodology, from the underlying molecular engineering paradigm (e.g., PCR) and associated cost/performance trade offs, to population sample selection consistent with societal norms and statistical needs, and safe (i.e., not risking further infections) operation of testing sites. Furthermore, in preparation for the day when more is known about the immune response to COVID-19 and possible vaccines, we plan to revise our models for feedback derived from a reliable immunoassay with well-specified performance parameters, such as lag times with respect to infection.

The paper is organized as follows. We summarize and explain the key findings in simple terms in Sec. II. In Sec. III, we discuss the use of massive testing as a direct means to contain the pandemics, showing that it requires a 100-fold increase of the current testing frequency. In Sec. IV, we define the main challenge to be addressed: To measure the quantitative effect of restrictive measures on the transmission rate. Section V introduces the idea of randomized testing. Section VI constitutes the central part of the paper, showing how data from sparse sampling tests can be used to infer essentially instantaneous growth rates, and their regional dependence. We define a model of policy interventions informed by feed-back from random testing and analyze it theoretically. The model is also analyzed numerically in Sec. VII. In Sec. VIII, we generalize the model for regionally refined analysis of the epidemic growth pattern which becomes the preferred choice if higher testing rates become available. We conclude with Sec. IX by summarizing our results and their implication for a safe reboot after the current lockdown. In the Supplementary Information, we address contact tracing and argue that it can complement, but not substitute for random testing. Finally, we present the algorithm used for our numerical results.

## II. SUMMARY OF KEY RESULTS

We argue that the moderate number of 15’000 random tests per day yields valuable information on the dynamics of the disease. Assuming that at a given time a conservatively estimated fraction of about *i*^*\**^ ≈ 0.07% of the population is currently infected [see Eq. 15d], on the order of 10 infected people will be detected every day. Can such a small number of detected infections be useful at all, given that these numbers fluctuate significantly from day to day? The answer is yes. We show that after a few days the acquired signal becomes stronger than the noise level. It is then possible to establish whether the infection number is growing or decreasing and, moreover, to obtain a quantitative estimate of the instantaneous growth rate *k*(*t*).

One of our central results is Eq. (13a) for the time where the signal becomes clear, which we rewrite in the simplified form

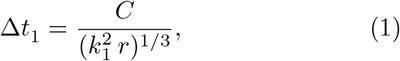

where *k*_1_ is the current growth rate of infections to be detected, and *r* is the number of tests per day ^3^. The numerical constant *C* depends on the required signal to noise ratio. A typical value when detecting large values of *k*_1_ is *C* ≈ 30 − 40.

This result shows that the higher the number of tests *r* per day, the shorter the time to detect a growth or a decrease of the infected population. The smaller the current growth rate *k*_1_, the longer the time to detect it above the noise inherent to the finite sampling.

How long would it take to detect that a release of restrictive measures has resulted in a nearly unmitigated growth rate of the order of *k*_1_ = 0.23 (which corresponds to doubling every 3 days)? Even with a moderate number of *r* = 15′000 per day, we find that within only Δ*t*_1 ≈_3 − 4 days such a strong growth will emerge above the noise level, such that countermeasures can be taken (see Fig. 6). During this short time, the damage remains limited. The infection numbers will have risen by a multiplicative factor between 2 and 3. This degree of control must be compared to a situation where no information on the current growth rate is available, and where the first effects of a new policy are seen in the increased number of symptomatic, sick people only 10-14 days later. Over this time span, with a growth rate of *k*_1_ = 0.23, the infection numbers will have grown by a factor of 10-30 before one realizes eventually that an intervention must be made.

Random testing decreases both the time scale until informed policy adjustments can be taken and the temporal fluctuations of the infection numbers. As in any feedback and control loop, the more frequent the testing is, the shorter are the delay times, and thus the smaller are the fluctuations. The various benefits of increasing the testing frequency are shown in Fig. 5, which are obtained by simulating a specific mitigation strategy, where we built in the uncertainty about the efficacy of political interventions. The shorter delay times and the reduced fluctuations result in decreased strain on the health system, lower economic costs, and a lower number of required interventions.

In addition to these benefits, a higher testing rate *r* also opens the opportunity to analyze geographic differences and refine the mitigation strategy accordingly, as we discuss in Sec. VIII.

## III. MASSIVE TESTING

If the massive frequency of 1.5 million tests per day becomes available in Switzerland, it will be possible to test any Swiss resident every 5 to 6 days. If the infected people that have been detected are kept in strict quarantine (such that they will not infect anybody anymore with high probability), such massive testing could be sufficient to prevent an exponential growth in the number of cumulated infections without the need of draconian physical distancing measures. We now explain qualitatively our approach to reach this conclusion (Ref. [5] gives a more detailed quantitative analysis).

The required testing rate can be estimated as follows. Let Δ*T* denote the average time until an infected person infects somebody else. The reproduction number *R*, i.e., the number of new infections transmitted on average by an infected person, falls below 1 (and thus below the threshold for exponential growth) if non-diagnosed people are tested at time intervals of no more than 2Δ*T*. Thus, the required number of tests over the time 2Δ*T*, the full testing rate 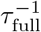, is

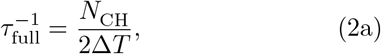

where

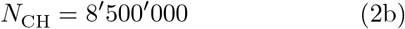

is the number of inhabitants of Switzerland. ^4^ Without social restrictions, it is estimated that [9]

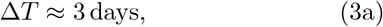

such that

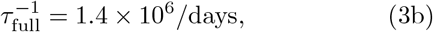

i.e., about 1.4 million tests per day would be required to control the pandemics by testing only. If additional restrictions such as physical distancing etc., are imposed, Δ*T* increases by a modest factor and one can get by with indirectly proportionally fewer tests per day. Nevertheless, on the order of 1 million tests per day is a minimal requirement for massive testing to contain the pandemics without further measures.

However, even while the Swiss capabilities are still far from reaching 1 million tests per day, testing for infections offers two important benefits in addition to identifying people that need to be quarantined. First, properly randomized testing allows to monitor and study the efficiency of measures that keep the reproduction number *R* below 1. This ensures that the growth rate *k* of case numbers and new infections is negative, *k* < 0. Second, frequent testing, even if applied to randomly selected people, helps suppress the reproduction number *R* and thus allows policy to be less restrictive in terms of other measures, such as physical distancing.

To quantify the latter benefit, observe that the effect of massive testing on the growth rate *k* is proportional to the testing rate [5]. Let us assume that without testing or social measures one has a growth rate *k*_0_. Then, if the testing rate 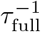 is sufficient to completely suppress the exponential growth in the absence of other measures, a smaller testing rate *τ*^−1^ decreases the growth rate *k*_0_ down to 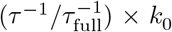. The remaining reduction of *k* to zero must then be achieved by a combination of restrictive social measures and contact tracing.

It is possible to refine the argument above to take account of the possibility of a spectrum of tests with particular cost/performance trade offs, i.e., a cheaper test with more false negatives could be used for random testing, whereas those displaying symptoms would be subjected to a “gold standard” (PCR) assay of viral genetic material.

## IV. QUANTIFYING THE EFFECTIVENESS OF RESTRICTIONS

A central challenge for establishing reliable predictions for the time evolution of a pandemic is the quantification of the effect of social restrictions on the transmission rate [3]. Policymakers and epidemiologists urgently need to know by how much specific restrictive measures reduce the growth rate *k*. Without that knowledge, it is essentially impossible to take an informed decision on how to optimally combine such measures to achieve a (marginally) stable situation, defined by the condition of a vanishing growth rate

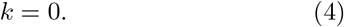

Indeed, marginal stability is optimal for two reasons. First, it is sustainable in the sense that the burden on the health system does not grow with time. Second, it is the least economically and socially restrictive state compatible with the stability requirement.

In Secs. V and VI, we suggest how marginal stability can be achieved, while simultaneously measuring the effects of a particular set of restrictions.

## V. STATISTICAL TESTING

We claim that statistically randomized testing can be used in a smart way, so as to keep the dynamics of the pandemics under control as per the feedback loop of Fig. 1. We emphasize that this is possible without the current time delays of up to 14 days. The latter arises since we only observe confirmed infections stemming from a highly biased test group that eventually shows symptoms long after the initial infection has occurred.

The idea of statistical testing is to *randomly* select people ^5^ and test them for infectiousness. ^6^

We stress that randomized testing is essential to obtain information on the current number of infections and its evolution with time. It serves an additional and entirely different purpose from testing people with symptoms, medical staff, or people close to somebody who has been infected, all of whom constitute highly biased groups of people.

The first goal of random testing is to obtain a firm test/confirmation of whether the current restrictive measures are sufficient to mitigate or suppress the exponential growth of the COVID-19 pandemic, ^7^ and whether the effectiveness differs from region to region. In case the measures should still be insufficient, one can measure the current growth rates and monitor the effect of additional restrictive measures.

## VI. NATIONAL MODELING AND INTERVENTION

We first analyze random testing for the case where we treat the country as a single entity with a population *N*. This will allow us to understand how testing frequency affects key characteristics of policy strategies.

### A. Model assumptions

We consider a model with the following idealizing assumptions:

1. An unbiased representative sample of the population is tested. A bias may underestimate the most relevant growth rate.
2. The rate of false positive tests is much less than the expected frequency of detection of infections.
3. Tests show whether a person is acutely infected in a short time (on the order of one day).
4. Policy measures can be applied rapidly, and their effect is immediate. Time delays due to the adaptation of human behavior to new rules is neglected.
5. The population is homogeneous as far as interactions between its members are concerned, e.g., there are no (semi)-isolated subpopulations. We do not account for large deviations in infectiousness that may lead to superspreading events [10].

As is well known to epidemiologists and the medical profession, the assumptions (1-4) clearly are violated to varying degrees in reality, but they can be taken into account by refinements of our model, whose operating principles and basic behavior will remain qualitatively the same. On the other hand, violations of assumption (5) can lead to new and dangerous effects, namely hotspots related to Anderson localization [11], which we discuss in Sec. VIII. Let *U* be the actual number of currently infected, but yet undetected people. (As in Ref. [5], we assume that detected people do not spread the disease.) The spreading of infections is assumed to be governed by the inhomogeneous, linear growth equation

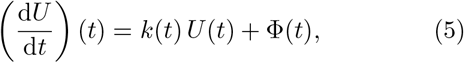

where *k*(*t*) is the instantaneous growth rate and Φ(*t*) accounts for infections arising from people crossing the national border. For simplicity, we set this influx to zero in this paper, in which case 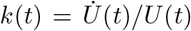 with the short-hand notation 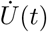 for the time derivative on the left-hand side of (5).

An equation of the form (5) is usually part of a more refined epidemiological model [12–14] that accounts explicitly for the recovery or death of infected persons. For our purpose, the effect of these has been lumped into an overall time-dependence of the rate *k*(*t*). For example, it evolves as the number of immune people grows, restrictive measures change, mobility is affected, new tracking systems are implemented, hospitals reach their capacity, testing is increased, etc. Nevertheless, over a short period of time where such conditions remain constant, and the fraction of immune people does not change significantly, we can assume the effective growth rate *k*(*t*) to be piece-wise constant in time. ^8^ We will exploit this below.

### B. Modeling intervention strategies

For *t* < 0, we assume a situation that is under control, with a negative growth rate

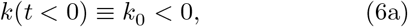

as is the case in Switzerland since the lockdown in March, with *k*_0_ ≈ − 0.07 day^−1^, according to the estimates of Ref. [4]. Such a stable state needs to be reached before a reboot of the economy can be considered. At *t* = 0 restrictive measures are first relaxed, resulting in an increase of the growth rate *k* from *k*_0_ to *k*_1_, which we assume positive,

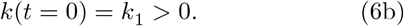

Hence, compensating counter measures are required at later times to avoid another exponential growth of the pandemic.

We now want to monitor the performance of policy strategies that relax or re-impose restrictions, step by step. The goal for an optimal policy is to reach a marginally stable state (4) (i.e., with *k* = 0) as smoothly, safely, and rapidly as possible. In other words, marginal stability is to be reached with the least possible damage to health, economy, and society. This expected outcome is to be optimized while controlling the risk of rare fluctuations.

To model the performance of policy strategies, we neglect the contributions to the time evolution of *k*(*t*) due to the increasing immunity or the evolution in the age distribution of infected people. We also neglect periodic temporal fluctuations of *k*(*t*) (e.g., due to alternation between workdays and weekends), which can be addressed in further elaborations. Instead, we assume that *k*(*t*) changes only in response to policy measures which are taken at specific times when certain criteria are met, as defined by a policy strategy. An intervention is made when the sampled testing data indicates that with high likelihood, *k*(*t*) exceeds some upper threshold

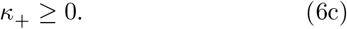

Likewise, a different intervention is made should *k*(*t*) be detected to fall below some negative threshold

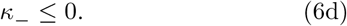

Note that if there is substantial infection influx Φ(*t*) across the national borders, one may want to choose the threshold *κ*_+_ to be negative, to avoid a too large response to the influx. From now on we neglect the influx of infections, and consider a homogeneous growth equation.

To reach decisions on policy measures, data is acquired by daily testing of random sets of people for infections. We assume that the tests are carried out at a limited rate *r* (a finite number of tests divided by a nonvanishing unit of time). Let *i*(*t*, Δ*t*) be the fraction of positive infections detected among the *r* Δ*t* ≫1 tests carried out in the time interval [*t, t* + Δ*t*]. By the law of large numbers, it is a Gaussian random variable with mean

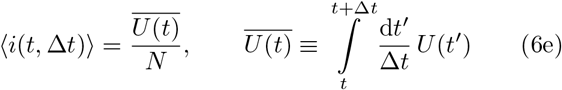

and standard deviation

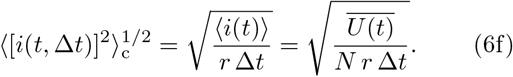

The current value of *k*(*t*) is estimated as *k*^fit^(*t*) by fitting these test data to an exponential, *where only data since the last policy change should be used*. The fitting also yields the statistical uncertainty (standard deviation), which we call *δk*(*t*). It will take at least 2-3 days to make a fit that is reasonably trustworthy.

If the instability threshold is surpassed by a certain level, i.e., if

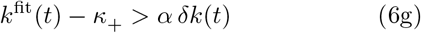

a new restrictive intervention is taken. If instead

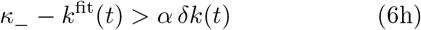

a new relaxing intervention is taken. Here, the parameter *α* is a key parameter defining the policy strategy. It determines the confidence level

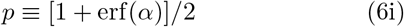

that policymakers require, before deciding to declare that a stability threshold has indeed been crossed. This strategy will result in a series of intervention times

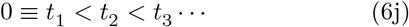

starting with the initial step to reboot at *t*_1_ = 0. In the time window [*t*_*ι*_, *t*_*ι*+1_], the growth rate *k*(*t*) is constant and takes the value

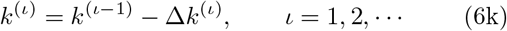

where a policy choice with Δ*k*^(*ι*)^ > 0 (corresponding to a restrictive measure) is made to bring back *k*(*t*) below the upper threshold *κ*_+_, while a policy choice with Δ*k*^(*ι*)^ < 0 is made to bring back *k*(*t*) above the lower threshold *κ*. The difficulty for policymakers is due to the fact that so far the quantitative effect of an intervention is not known. We model this uncertainty by assuming Δ*k*^(*ι*)^ to be random to a certain degree.

If at time *t, k*^fit^(*t*) crosses the upper threshold *κ*_+_ with confidence level *p*, we set *t*_*ι*_ = *t* and a restrictive measure is taken, i.e., Δ*k*^(*ι*)^ is chosen positive. We take the associated decrement Δ*k*^(*ι*)^ to be uniformly distributed on the interval

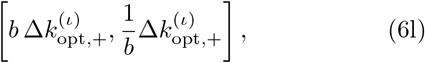

where the optimum choice 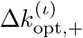 is defined by

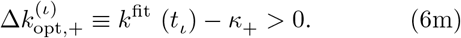

The parameter *b* < 1 describes the uncertainty about the effects of the measures taken by policymakers. While the policymakers aim to reset the growth factor *k* to *κ*_+_, the result of the measure taken may range from having an effect that is too small by a factor of *b* to overshooting by a factor of 1*/b*. A measure with effect 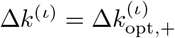 would be optimal according to the best current estimate. The larger 1 − *b*, the larger the uncertainty. Unless stated otherwise, we assume *b* = 0.5.

If instead *k*^fit^(*t*) crosses the lower threshold *κ*_−_ with confidence level *p* at time *t*, we set *t*_*ι*_ = *t* and a relaxing measure is taken, i.e., Δ*k*^(*ι*)^ is chosen negative. Again, Δ*k*^(*ι*)^ is uniformly distributed on the interval

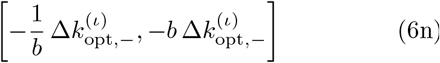

with the optimum choice 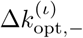 defined by

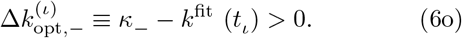

The process described above is stochastic for two reasons. First, the sampling comes with the usual uncertainties in the law of large numbers. Second, the effect of policy measures is not known beforehand (even though it may be learnt in the course of time, which we do not include here). It should be clear that the faster the testing the more rapidly one can respond to a super-critical situation.

A significant simplification of the model occurs when the two thresholds are chosen to vanish,

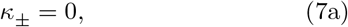

in which case

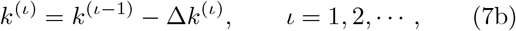

with |Δ*k*^(*ι*)^| uniformly distributed on the interval

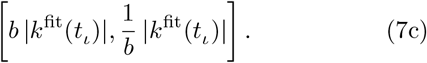

In this case the system will usually tend to a critical steady state with *k*(*t* →∞) → 0, as we will show explicitly below. In this case the policy strategy can simply be rephrased as follows. As soon as one has sufficient confidence that *k* has a definite sign, one intervenes, trying to bring *k* back to zero. The only parameter defining the strategy is *α*.

### C. Testing and fitting procedure

Let us now detail the fitting procedure and analyze the typical time scales involved between subsequent policy interventions when choosing the thresholds (7). After a policy change at time *t*_*ι*_, data is acquired over a time window Δ*t*. We then proceed with the following steps to estimate the time *t*_*ι*+1_ at which the next policy change must be implemented.

*Step 1: Measurement* We split the time window

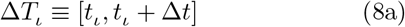

of length Δ*t* after the policy change into the time interval

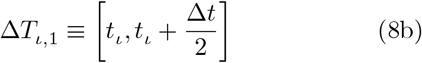

and the time interval

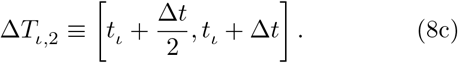

Testing delivers the number of currently infected people

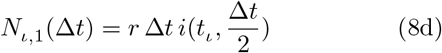

for the time interval (8b) and

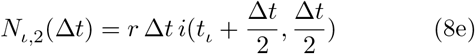

for the time interval (8c), where we recall that *r* denotes the number of people tested per unit time. Given those two measurements over the time window Δ*t/*2, we obtain the estimate

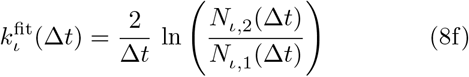

with the standard deviation

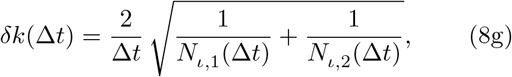

as follows from the statistical uncertainty 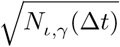 of the sampled numbers *N*_*ι,γ*_ (Δ*t*) and standard error propagation. The above recipe can be replaced by a more sophisticated Levenberg-Marquardt fitting procedure, which yields more accurate estimates for *k*(*t*) with a smaller uncertainty *δk*(*t*). We have confirmed that this uniformly improves the performance of the mitigation strategy.

*Step 2: Condition for new policy intervention* A new policy intervention is taken once the magnitude 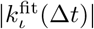 with 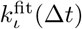 given by Eq. (8f) exceeds *α δk*(Δ*t*) with *δk*(Δ*t*) given by Eq. (8g). Here, *α* controls the accuracy to which the actual *k* has been estimated at the time of the next intervention. The condition

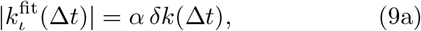

for a new policy intervention thus becomes

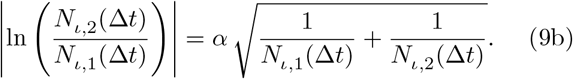

*Step 3: Comparison with modeling* We call *i*(*t*) = *U* (*t*)*/N* the actual fraction of infections (in the entire population) as a function of time, which we assume to follow a simple exponential evolution between two successive policy interventions, i.e., the normalized solution

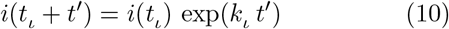

to the growth equation (5) on the interval *t*_*ι*_ < *′* < *t*_*ι*+1_. The expected number of newly detected infected people in the time interval (8b) is

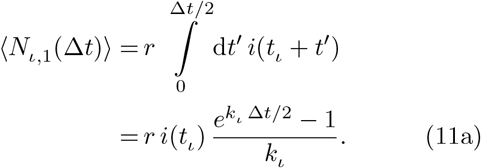

Similarly, the predicted number of infected people in the time interval (8c) is

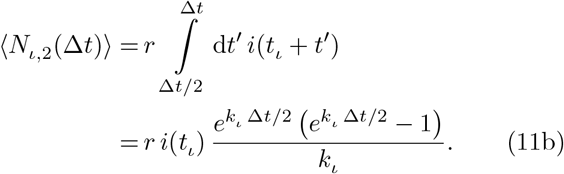

*Step 4: Estimated time for a new policy intervention* We now approximate *N*_*ι*,1_ and *N*_*ι*, 2_ by replacing them with their expectation value Eqs. (11a) and (11b), respectively, and anticipating the limit

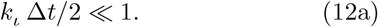

We further anticipate that for safe strategies the fraction of currently infected people *i*(*t*) does not vary strongly over time. More precisely, it hovers around the value *i*^*\**^ defined in Eqs. (14b) and (15d) (see Fig. 2). We thus insert

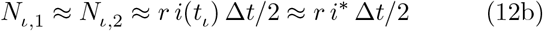

into Eq. (9b) and solve for Δ*t*. The solution is the time until the next intervention

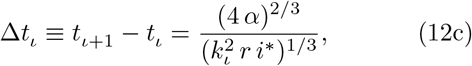

from which we deduce the relative increase

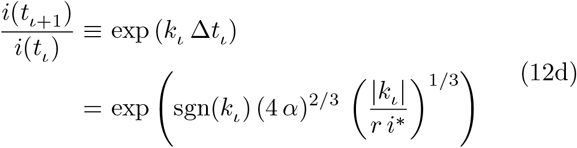

of the fraction of currently infected people over the time window. This relative increase is close to 1 if the argument of the exponential on the right-hand side is small.

We will show below that the characteristics

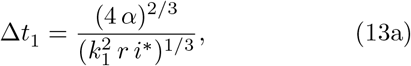

and

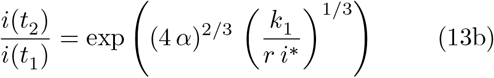

of the first time interval [*t, t*] set the relevant scales for the entire process. From Eqs. (12c) and (12d), we infer the following important result. The higher the testing frequency *r*, the smaller the typical variations in the fraction of currently infected people, and thus in the case numbers. The band width of fluctuations decreases as *r*^−1*/*3^ with the testing rate.

#### 1. Critical fraction of infections

As one should expect, it is always the average rate to detect a currently infected person, *r i*^*\**^, which enters into the expressions (12c) and (12d). The higher the fraction *i*^*\**^, the more reliable is the sampling, the shorter is the time to converge toward the marginal state (4), and the smaller are the fluctuations of the fraction of infected people.

If the fraction *i*^*\**^ is too low the statistical fluctuations become too large and little statistically meaningful information can be obtained. On the other hand, if the fraction of infections drops to much lower values, then policy can be considered to have been successful and can be maintained until further tests show otherwise.

We seek an upper bound for a manageable *i*^*\**^. Here we consider the parameters of Switzerland. However, they can easily be adapted to any other country. We assume that a fraction 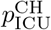 of infected people in Switzerland needs to be in intensive care. ^9^ Here, we will use the value 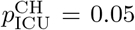. Let *ρ*_ICU_ be the number of ICU beds per inhabitant that shall be allocated to COVID-19 patients. The Swiss national average is about [15]

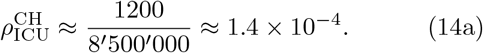

For the pandemics not to overwhelm the health system, one thus needs to maintain the fraction of currently infected people safely below

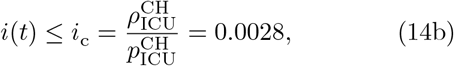

together with similar constraints related to the capacity for hospitalizations, medical care personnel and equipment for specialized treatments. We take the constraint from intensive care units to obtain an order of magnitude for the upper limit admissible for the infected fraction of people, *i*. A recent study based on random testing reports that the fraction of people currently infected with the virus lies within the confidence interval [0.0012, 0.0076] in Austria (whereby half of the infected people in the sample were previously undetected) [16]. The estimates in Ref. [4] suggest that the fraction of acutely infected people was even close to 0.01 before the lock-down in Switzerland. This indicates that our threshold estimate (14b) is conservative. If the actual threshold (which depends on the country, the structure of its population, and its health-care infrastructure) is higher, the testing frequency required to reach a defined accuracy decreases in proportion.

The objective is to mitigate the pandemic so that values of the order of *i*_c_ or below are achieved. Before that level is reached restrictions cannot be relaxed. It may prove difficult to push the fraction of infected people significantly below *i*_*c*_, since the recent experience in most European countries suggests that it is very hard to ensure that growth rates *k* fall well below 0. The main aim would then be to reach at least stabilization of the number of currently infected people (*k* = 0).

For the following we thus assume that the fraction of infections *i* will stagnate around a value *i*^*\**^ of the order of *i*_c_. We will discuss below what ratio *i*^*\**^*/i*_c_ can be considered safe.

#### D. Required testing rate

We seek the testing rate that is needed to obtain a strategy with satisfactory outcome. We assume that after the reboot at *t*_1_ = 0, the initial growth rate may turn out to be fairly high, say of the order of the unmitigated growth rate. In many European countries a doubling of cases was observed every three days before restrictive measures were introduced. This corresponds to a growth rate of

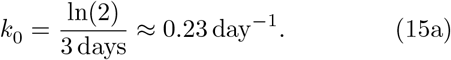

We assume an initial growth rate of

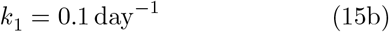

just after the reboot. We choose the reasonably stable confidence parameter

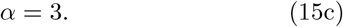

In Sec. VII we will find that this choice strikes a good balance between several performance criteria (see Fig. 4). We further assume that the rate of infections initially stagnates at a level of (for Switzerland)

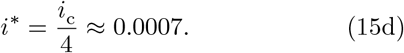

The level *i*^*\**^ should, however, be measured by random testing before a reboot is attempted. We should then ensure that the first relative increase of

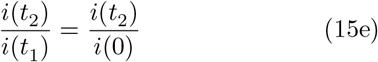

does not exceed a factor of 4. From Eq. (13b), we thus obtain the requirement

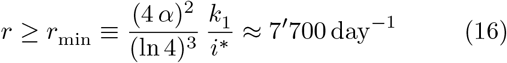

for the testing rate *r*. Note the inverse proportionality to the parameter *i*^*\**^, for which Eq. (15d) is a conservative estimate. Using this value yields an estimate of the order of magnitude required for Switzerland. In the next section we simulate a full mitigation strategy and confirm that with additional capacity for just about 15’000 random infection tests per day a nation-wide, safe reboot can be envisioned for Switzerland.

We close with two observations. First, this minimal testing frequency is just twice the testing frequency presently available for suspected infections and medical staff in Switzerland. Second, while the latter tests require a high sensitivity with as few false negatives as possible, random testing can very well be carried out with tests of lower quality in that respect. Indeed, an increase in false negatives acts as a systematic error in the estimate of the infected fraction of people, which, however, drops out in the determination of its growth rate, ^10^ as long as the fraction *i* is not close to 1. However, the success of random testing does rely on a very low probability (≪*i*^*\**^) of false positives (as is the case of current PCR tests). Otherwise the signal from true positives would rapidly be overwhelmed by the noise from false positives.

### E. Further intervention steps after the reboot

After the reboot at time *t*_1_ = 0 further interventions will be necessary, as we assume that the reboot will have resulted in a positive growth rate *k*_1_. In subsequent interventions, the policymakers try to take measures that aim at reducing the growth rate to zero. Even if they had perfect knowledge of the current growth rate *k*(*t*), they would not succeed immediately since they do not know the precise quantitative effect of the measures they will take. Nevertheless, had they complete knowledge of *k*(*t*), our model assumes that they would be able to gauge their intervention such that the actual effect on *k*(*t*) differs at most by a factor between *b* and 1*/b* from the targeted value, which would reduce *k*(*t*) to 0. This and the assumption *b* ≥ 0.5 implies that, if *α* is large, so that *k*(*t*) is known with relatively high precision at the time of intervention, the growth rate *k*_2_ is smaller than *k*_1_ in magnitude with high probability (tending rapidly to 1 as *α* →∞). ^11^ The smaller *α* however, the more likely it becomes, that *k*(*t*) is overestimated, and an exaggerated corrective measure is taken, which may destabilize the system. In this context, we observe that the ratio

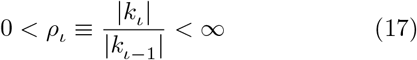

is a random variable with a distribution that is independent of *ι* in our model. To proceed, we assume that *α* is sufficiently large, such that the probability that *ρ*_ι_ < 1 is indeed high.

The second policy intervention occurs after a time that can be predicted along the same lines that lead to Eq. (12c). One finds

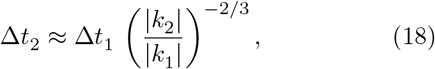

where Δ*t*_1_ is given by Eq. (13a). Since, the growth rate *k*_3_ is likely to be smaller than *k*_2_ in magnitude, the third intervention takes place after yet a longer time span, etc. If we neglect that the fitted value 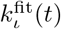 differs slightly from *k*_*ι*_ (a difference that is negligible when *α ≫* 1), our model ensures that *k*_*ι*_*/k*_*ι*1_ is uniformly distributed in [*b* − 1, 1*/b* − 1]. After the *ι*-th intervention the growth rate is down in magnitude to

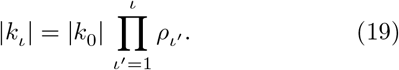

To reach a low final growth rate *k*_final_, a typical number *n*_int_(*k*_final_) of interventions are required after the reboot, where

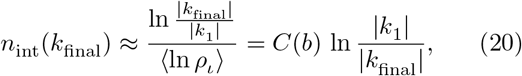

where the constant *C*(*b*) = − 1*/* ⟨ln *ρ*_*ι*_ ⟩ depends on the policy uncertainty parameter *b*.

The time to reach this low rate is dominated by the last time interval which yields the estimate

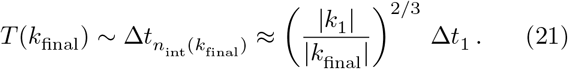

Thus, the system converges to the critical state where *k* = 0, but never quite reaches it. At late times *T*, the residual growth rate behaves as *k*_final_ ∼ *T* ^−3*/*2^.

### F. Choosing an optimal intervention strategy

The parameter *α* encodes the confidence policymakers need about the present state before they take a decision. Here we discuss various measures that allow choosing an optimal value for *α*.

As *α* decreases starting from large values, the time for interventions decreases, being proportional to *α*^2*/*3^ according to Eq. (13a). Likewise the fluctuations of infection numbers will initially decrease. However, the logarithmic average − ⟨ln *ρ*_*ι*_ ⟩ in the denominator of Eq. (20) will also decrease, and thus the necessary number of interventions increases. Moreover, when *α* falls below 1, interventions become more and more ill-informed and erratic. It is not even obvious anymore that the marginally stable state is still approached asymptotically. From these two limiting considerations, we expect

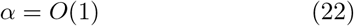

to be an optimal choice for *α*.

Let us now discuss a few quantitative measures of the performance of various strategies, which will allow policymakers to make an optimal choice of confidence parameter for the definition of a mitigation strategy.

#### 1. Time scale to approach the marginal state

The time to reach a certain level of quiescence (low growth rates, infrequent interventions) is given by the time (21), and thus by the expectation value of Δ*t*_1_.

#### 2. Political cost

As a measure for the political cost, *C*_P_, we may consider the number of interventions that have to be taken to reach quiescence. As we saw in Eq. (20), it scales inversely with the logarithmic average of the ratios of growth rates, *ρ*, i.e.,

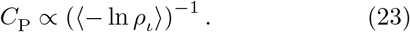

#### 3. Health cost

If restrictions are over-relaxed, the infection numbers will grow with time. The maximal fraction of currently infected people must never be allowed to rise above the manageable threshold of *i*_c_. This means that continuous (random) monitoring of the fraction of infected people is needed, so that given the knowledge from the time before the reboot, about the conditions under which the system can be stabilized, lockdown conditions can always be imposed at a time that is sufficient to prevent reaching the level of *i*_c_. Beyond this consideration one may want to keep the expected maximal increase of infection numbers low, which we take as a measure of health costs *C*_H_,

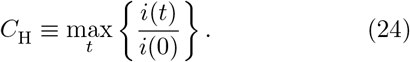

Note that as defined, *C*_H_ is a stochastic number. Its mean and tail distribution (for large *R*) will be of particular importance.

#### 4. Economic and social cost

Imposing restrictions such that *k* < 0 imply restrictions beyond what is absolutely necessary to maintain stability. If we assume that the economic cost *C*_E_ is proportional to the excess negative growth rate, − *k* (and a potential gain proportional to −*k*), a measure for the economic cost is the summation over time of −*k*(*t*),

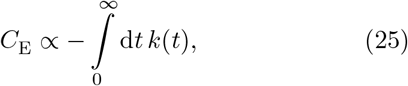

which converges, since *k*(*t*) decays as a sufficiently fast power law. Hereto, *C*_E_ is a stochastic variable that depends on the testing history and the policy measures taken. However, its mean and standard deviation could be used as indicators of economic performance.

## VII. SIMULATION OF MITIGATION STRATEGY BY RANDOM TESTING

We introduced in Sec. VI a feedback and control strategy to tune to a marginal state with vanishing growth rate *k* = 0 after an initial reboot. Interventions were only taken based on the measurement of the growth rate. However, in practice, a more refined strategy will be needed. In case the infection rate drops significantly below *i*^*\**^, one might (depending on netting out political and economic pressures, something which the authors of this paper are not doing here) benefit from a positive growth rate *k*. We thus assume that if *i*(*t*)*/i*^*\**^ falls below some threshold *i*_low_ = 0.2, we intervene by relaxing some measures, that we assume to increase *k* by an amount uniformly distributed in [0, *k*_1_], but without letting *k* exceed the maximal value of *k*_high_ = 0.23. Likewise, one should intervene when the fraction *i*(*t*) grows too large. We do so when *i*(*t*)*/i*^*\**^ exceeds *i*_high_ = 3. In such a situation we impose restrictions resulting in a decrease of *k* by a quantity uniformly drawn from [*k*_high_*/*2, *k*_high_]. The precise algorithm is given in the Supplementary Information.

Figure 3 shows how our algorithm implements policy releases and restrictions in response to test data. The initial infected fraction and growth rate are *i*(0) = *i*_c_*/*4 = 0.0007 and *k*_1_ = 0.1, respectively, with a sampling interval of one day. We choose *α* = 3 and a number of *r* = 15^*/*^000 tests per day. Figure 3(a) displays the infection fraction, *U* (*t*)*/N*, as a function of time, derived using our simple exponential growth model, which is characterized by a single growth rate that changes stochastically at interventions [Eq. (5) without the source term]. In the absence of intervention, the infected population would grow rapidly representing uncontrolled runaway of a second epidemic. At each time step (day) the currently infected fraction of the population is sampled. The result is normally distribution with mean and standard deviation given by Eqs. (6e) and (6f) to obtain *i*(*t*). The former are represented by small circles, the latter by vertical error bars in Fig. 3. If *i/i*^*\**^ lies outside the range [*i*_low_, *i*_high_], we intervene as described above. Otherwise, on each day *k*^fit^(*t*) and its standard deviation are estimated using the data since the last intervention. With this, at each time step, Eqs. (6m) to (6o) decide whether or not to intervene. In Fig. 3, each red circle represents an intervention and therefore either a decrease or increase of the growth rate constant of our model.

**FIG. 3.**
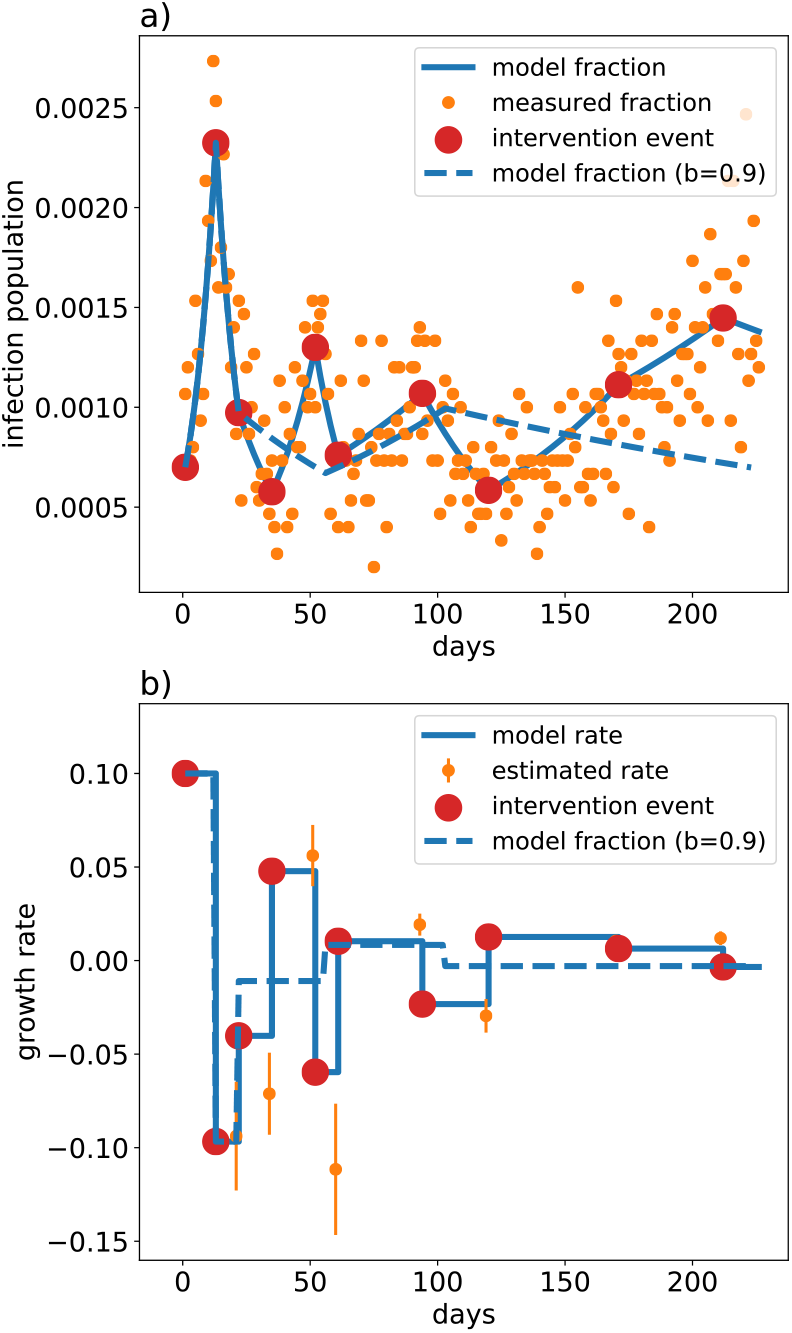
Our algorithm implements policy releases and restrictions aiming at maintaining a vanishing growth rate. It intervenes whenever the estimated slope of the fraction of infected people is found to be non-zero, here with confidence level *α* = 3. We plot the model infection fraction *U* (*t*)*/N* and the detected infection fraction *i*(*t*) as a function of days in panel (a). The model growth rate *k*(*t*) (solid line) and the estimated growth rate *k*_est_ at times of intervention are shown in panel (b) for the parameters *i*(0) = 0.0012, *k*_1_ = 0.1, and a test rate of *r* = 15′000 day^−1^. The dashed blue line corresponds to a history of interventions where we assumed that the effect of policy interventions is better known (described by an uncertainty parameter *b* = 0.9, instead of *b* = 0.5), so that convergence is much faster.

Figure 3 shows the evolution of the fraction of currently infected people. After an initial growth with rate *k*_1_ subsequent interventions reduce the growth rate down to low levels within a few weeks. At the same time the fraction of infected people stabilizes at a scale similar to *i*^*\**^. For the given parameter-set this is a general trend independent of realization. Figure 3(b) displays the instantaneous value of the model rate constant and also the estimated value together with its fitting uncertainty. The estimate follows the model value reasonably well. One sees that the interventions occur when the uncertainty in *k* is sufficiently small.

### A. Simulation results

We now assume that we have the capacity for *r* = 15^*/*^000 per day, and assess the performance of our strategy as a function of the confidence parameter *α* in Fig. 4. Values of *α* ≤2 lead to rapid, but at the same time erratic interventions, as is reflected by a rapidly growing number of interventions. For larger values of *α*, the time scale to reach a steady state increases while the economic and health costs remain more or less stable. A reasonable compromise between minimizing the number of interventions, and shortening the time to reach a steady state suggests a choice of *α* ≈ 2.5 − 3.5.

**FIG. 4.**
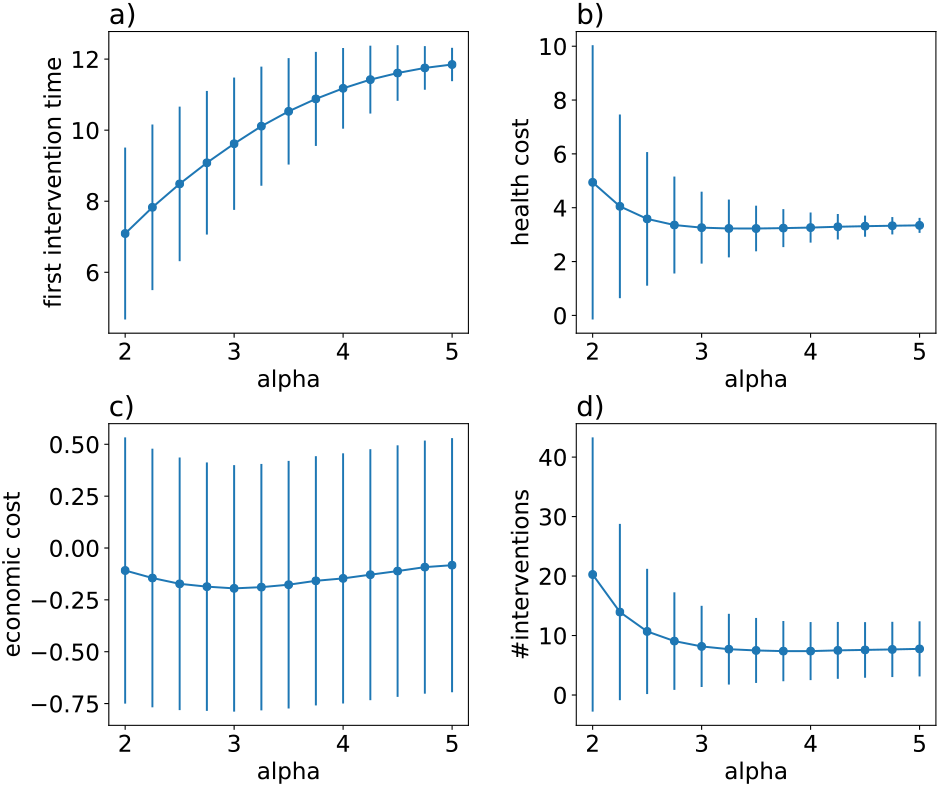
Performance of the mitigation strategy as a function of the confidence parameter *α*, for a number *r* = 15 ′000 tests per day and an initial growth rate *k*_1_ = 0.1. We plot the time scale Δ*t*_1_ (a), and the health (b), economic (c) and political (as measured by numbers of interventions to achieve a steady state) (d) costs [Eqs. (23)–(25)] as measures of performance. The circles are the mean values, the vertical lines indicate the standard deviations of the respective quantities.

It is intuitive that the higher the number *r* of tests per day is, the better the mitigation strategy will perform. The characteristic time to reach a final steady state decreases as *r*^−1*/*3^, see Eq. (13a). Other measures for performance improve monotonically upon increasing *r*. This is confirmed and quantified in Fig. 5, where we show how the political, health, and economic cost decreases with increasing test rate.

**FIG. 5.**
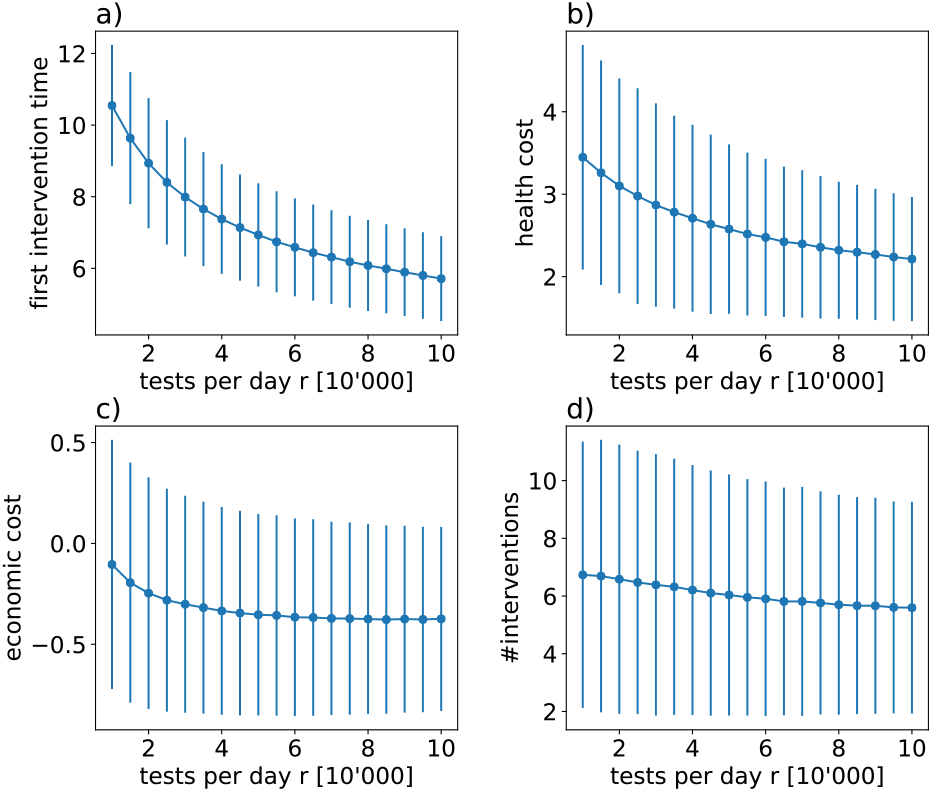
Performance of the mitigation strategy as a function of the number of tests *r* per day, for a fixed value of *α* = 3 and an initial growth rate *k*_1_ = 0.1. We plot the time scale Δ*t*_1_ (a), and the health (b), economic (c) and political (as measured by numbers of interventions to achieve a steady state (d) costs [Eqs. (23)–(25)] as measures of performance. The circles are the mean values, the vertical lines indicate the standard deviations of the respective quantities. The large uncertainties in the economic costs, e.g., are a consequence of the relatively large uncertainty in the effect of interventions (*b* = 0.5). If the latter is better known, the standard deviation of the cost functions will decrease accordingly.

#### 1. Time delay to detect catastrophic growth rates

After a reboot it is likely that the growth rate *k*_1_ jumps back to positive values, as we have always assumed so far. The time it takes until one can distinguish a genuine growth from intrinsic fluctuations due to the finite number of sample people depends on the growth rate *k*_1_, see Eq. (13a).

In the worst case where the reboot brings back the unmitigated value *k*_0_, one will know within 3-4 days with reasonable confidence that the growth rate is well above zero. This is shown in Fig. 6. In such a catastrophic situation, an early intervention can be taken, while the number of infections has at most tripled at worst. Note that this reaction time is 4-5 times faster than without random testing.

**FIG. 6.**
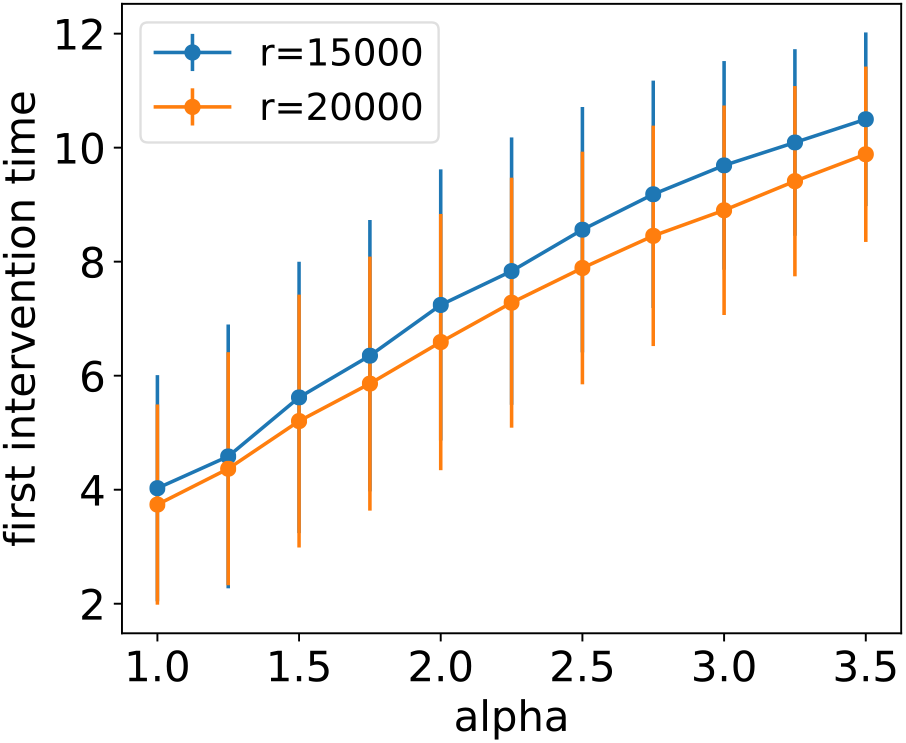
Time after which a significant positive growth rate is confirmed in the worst case scenario for which the growth rate jumps to *k*_1_ = 0.23 after reboot. An intervention will be triggered in 3-4 days, since in the case that such a strong growth must be suspected, one should apply a small confidence parameter *α* ≈ 1. Results are shown for *r* = 15′000 and *r* = 20′000 tests a day. The circles are the mean values, the vertical lines indicate the standard deviations for the first intervention time.

## VIII. REGIONALLY REFINED REBOOT AND MITIGATION STRATEGIES

We have argued that the minimal testing rate *r*_min_ (16) is sufficient to obtain statistical information on the growth rate *k* as applied to Switzerland as a whole. This assumes tacitly that the simple growth equation (5) describes the dynamics of infections in the whole country well. That this is not necessarily a good description can be conjectured from recent data on the current rates with which numbers of confirmed infections in the various cantons (states of Switzerland) evolve. These data indeed show a very significant spread by nearly a factor of four suggesting that a spatially resolved approach is preferable, if possible.

If the testing capacity is limited by rates of order *r*_min_, the approach can still be used. But caution should be taken to account for spatial fluctuations corresponding to hot spots. One should preferentially test in areas that are likely to show the largest local growth rates so as not to miss locally super-critical growth rates by averaging over the entire country. If however, higher testing frequencies become available, new and better options come into play.

### A. Partitioning the country for statistical analysis

Valuable information can be gained by analyzing the test data not only for Switzerland as a whole, but by distinguishing different regions.

It might even prove useful not to lift restrictions homogeneously throughout the country, but instead to vary the set of restrictions to be released, or to adapt their rigor. By way of example, consider that after the spring vacation school starts in different weeks in different cantons. This regional difference could be exploited to probe the relative effect of re-opening schools on the local growth rates *k*. However, obviously, it might prove politically difficult to go beyond such “naturally” occurring differences, as it is a complex matter to decide what region releases which measures first. A further issue is that the effects might be unclear at the borders between regions with different restrictions. There may also be complications with commuters that cross regional borders. Finally, there may be undesired behavioral effects, if regionally varying measures are declared as an “experiment”. Such issues demand careful consideration if regionally varying policies are applied.

Even if policy measures should eventually not be taken in a region-specific manner, it is very useful to study a regionally refined model of epidemic dynamics. Indeed a host of literature exists that studies epidemiological models on lattices and analyzes the spatial heterogeneities. [17, 18] In certain circumstances those have been argued to become even extremely strong. [19] In the present paper, we will content ourselves with a few general remarks concerning such refinements. We reserve a more thorough study of regionally refined testing and mitigation strategies to a later publication.

Let us thus group the population of Switzerland into *G* sets. The most natural clustering is according to the place where people live, cities or counties. ^12^ The more we partition the country, the more spatially refined the acquired data will be, and the better tailored mitigation strategies could potentially become. However, this comes at a price. Namely, for a limited national testing rate *r*_tot_, an increased partitioning means that the statistical uncertainty to measure local growth rates in each region will increase.

The minimal test rate *r*_min_ that we estimated on the right-hand side of Eq. (16) still holds, but now for each region, which can only test at a rate *r* = *r*_tot_*/G*. To refine Switzerland *G* regions we thus have the constraint that the total testing capacity exceeds

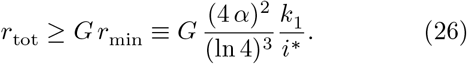

On the other hand, if testing at a high daily rate *r*_tot_ becomes available, nothing should stop one to refine the statistical analysis to *G* ≈ *r*_tot_*/r*_min_ to make the best use of available data.

### B. Spatially resolved growth model

Each of the population groups *m* ∈ {1,, *G*}is assumed to have roughly the same size, containing

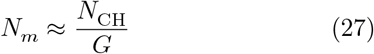

people, *U*_*m*_ of whom are currently infected, but yet undetected. The spreading of infections is again assumed to follow a linear growth equation (where we neglect influx from across the borders from the outset)

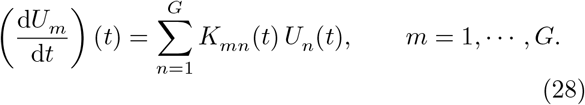

Here, the growth kernel *K*(*t*) is a *G* × *G* matrix with matrix elements *K*_*mn*_(*t*). The matrix *K*(*t*) has *G* (complex valued) eigenvalues *λ*_*n*_, *n* = 1,, *G*. The largest growth rate is given by

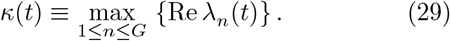

For the sake of stability criteria, *κ*(*t*) now essentially takes the role of *k*(*t*) in the model with a single region, *G* = 1. We note that the number of infections grows exponentially if *κ*(*t*) > 0, and decreases if *κ*(*t*) < 0.

As in the case of a single region, we assume *K*(*t*) to be piece wise constant in time, and to change only upon taking policy interventions.

In the simplest approximation, one assumes no contact between geographically distinct groups, that is, the offdiagonal matrix elements are set to zero [*K*_*m*≠*n*_(*t*) = 0] and the eigenvalues become equal to elements of the diagonal: *k*_*m*_(*t*) *≡ K*_*mm*_(*t*). As current cantonal data suggests, the local growth rate *k*_*m*_(*t*) depends on the region, and thus *k*_*m*_(*t*) *k*_*n*_(*t*). It is natural to expect that *k*_*m*_(*t*) correlates with the population density, the fraction of the population that commutes, the age distribution, etc.

If on top of the heterogeneity of growth rates one adds finite but weak inter-regional couplings _*Km* ≠*n*_(*t*) > 0 (mostly between nearest neighbor regions), one may still expect the eigenvectors of *K*(*t*) to be rather localized (a phenomenon well known as Anderson localization [11] in the context of waves propagating in strongly disordered media). By this, one means that the eigenvectors have a lot of weight on few regions only, and little weight everywhere else. That such a phenomenon might occur in the growth pattern of real epidemics is suggested by the significant regional differences in growth rates that we have mentioned above. In such a situation it would seem preferable to adapt restrictive measures to localized regions with strong overlap on unstable eigenvectors of *K*(*t*), while minimizing their socio-economic impact in other regions with lower *k*_*m*_(*t*).

### C. Mitigation strategies with regionally refined analysis

As mentioned above, in the case with several distinct regions, *G* > 1, an intervention becomes necessary when the largest eigenvalue *κ*(*t*) of *K*(*t*) crosses an upper or a lower threshold (with a level of confidence *α* again to be specified). If the associated eigenvector is delocalized over all regions, one will most likely respond with a global policy measure. However, it may as well happen that the eigenvector corresponding to *κ*(*t*) is well-localized. In this case one can distinguish two strategies for intervention:

a. **Global strategy** One always applies a single policy change to the whole country. This is politically simple to implement, but might incur unnecessary economic cost in regions that are not currently unstable.
b. **Local strategy** One applies a policy change only in regions which have significant weight on the unstable eigenvectors. This means that one only adjusts the corresponding diagonal matrix elements of *K*(*t*) and those off-diagonals that share an index with the unstable region.

Likewise, regions that have *i*_*m*_ < *i*^*\**^ and have negligible overlap with eigenvectors whose eigenvalues are above *κ*_−_, could relax some restrictions before others do.

Fitting test data to a regionally refined model will allow us to estimate the off-diagonal terms *K*_*mn*_(*t*), which are so far poorly characterized parameters. However, the *K*_*mn*_(*t*) contain valuable information. For instance, if a hot spot emerges [that is, a region overlapping strongly with a localized eigenvector with positive Re *λ*_*n*_(*t*)], this part of the matrix will inform which connections are the most likely to infect neighboring regions. They can then be addressed by appropriate policy measures and will be monitored subsequently, with the aim to contain the hot spot and keep it well localized.

This model allows us to calculate again economic, political, and health impact of various strategies. It is important to assess how the global and the local strategy perform in comparison. Obviously this will depend on the variability between the local growth rates *k*_*m*_(*t*), which is currently not well known, but will become a measurable quantity in the future. At that point one will be able to decide whether to select the politically simpler route (a) or the heterogeneous route (b) which is likely to be economically favorable.

We are currently engaged in developing an analysis tool to quickly process test data for multi-region modeling. We are developing and assessing intervention strategies with the perspective of running it daily with the best available current data and knowledge.

## IX. SUMMARY AND CONCLUSION

We have analyzed a feedback and control model for managing a pandemic such as that caused by COVID-19. The crucial output parameters are the infection growth rates in the general population and spatially localized sub-populations. When planning for an upcoming reboot of the economy, it is essential to assess and mitigate the risks of relaxing some of the restrictions that have brought the COVID-19 epidemic under control. In particular, the policy strategy chosen must suppress a potential second exponential wave when the economy is rebooted, and so avoid a perpetual stop-and-go oscillation between relaxation and lockdown. Feedback and control models are designed with precisely this goal in mind.

Having random testing in place, the risk of a second wave can be kept to a minimum. Additional testing capacity of *r* = 15′000 day^−1^ tests (on top of the current tests for medical purposes) carried out with randomly selected people would allow us to follow the course of the pandemics almost in real time, without huge time delays, and without the danger of increasing the number of currently infected people by more than a factor of four, if our intervention strategy is followed. We emphasize that our estimate of *r* is conservative. If the manageable fraction of infected people is higher than what we assumed in Eq. (14b), namely of order *i*_c_ ≈0.01 as the estimates of Ref. [4] suggest, the required testing rate decreases by a factor 3 − 4 to a mere *r* = 4′000 − 5′000 day^−1^.

If testing rates *r* significantly higher than *r*_min_ become available, a regionally refined analysis of the growth dynamics can be carried out, with *G* ≈*r/r*_min_ regions that can be distinguished.

In the worst case scenario, where releasing certain measures immediately make the country jump back to the unmitigated growth rate of *k*_0_ = 0.23 day^−1^, random testing would detect this within 3-4 days from the change coming into effect. This is in stark contrast to the nearly 14 days of delay required for symptomatic individuals to emerge in statistically significant numbers. After such a time delay a huge increase (by a factor of order 20) of infection numbers may have already occurred, which would be catastrophic. Daily random testing safely prevents this. Thereby the significant reduction of the time delay is absolutely crucial. Note that without daily polling of infection numbers and without knowledge about the quantitative effect of restriction measures, a reboot of the economy could not be risked before the number of infections has been suppressed by at least a factor of 10-20 below the current level. Given the limits of suppression rates that can be achieved without most draconic lock-down measures, this will require a very long time and thus translates into an enormous economic cost. In contrast, daily polling will allow us to carefully reboot the economy and adjust restrictive measures, while closely monitoring their effect. Since the reaction times are so much shorter, one can safely start an attempted reboot already at infection numbers corresponding roughly to the status quo.

At some point one might consider the option to start releasing different sets of restrictions in different regions, with the aim to learn faster about their respective effects and thus to optimize response strategies in subsequent steps.

## Data Availability

The data generated by the numerical simulations can be provided upon request to the authors.

## X. ACKNOWLEDGMENTS

We are grateful to Giulia Brunelli, Klaus Müeller, Emma Slack, Thomas Van Boeckel, and Li Zhiyuan for helpful discussions, and the ERC HERO project 810451 for supporting GA.

## Appendix A Assessment of contact tracing as a means to control the pandemics

Let us briefly discuss the strategy of so-called contact tracing as a means to contain the pandemics, as has been discussed in the literature [**?**]. We argue that contact tracing is a helpful tool to suppress transmission rates, but is susceptible to fail when no other method of control is used.

Contact tracing means that once an infected person is detected, people in their environment (i.e., known personal contacts, and those identified using mobile-phone based Apps etc) are notified and tested, and quarantined if detected positive. As a complementary measure to push down the transmission rate, it is definitely useful, and it represents a relatively low cost and targeted measure, since the probability to detect infected people is high. However, as a sole measure to contain a pandemic contact tracing is impractical (especially at the current high numbers of infected people) and even hazardous.

The reason is as follows. It is believed that a considerable fraction *f*_asym_ of infected people show only weak or no symptoms, so that they would not get tested under the present testing regime. The value of *f*_asym_ is not well known, but it might be rather high (30% or even much higher). Such asymptomatic people will go undetected, if they have not been in contact with a person displaying symptoms. If on average they infect *R* people while being infectious, and if *R f*_asym_ > 1, there will be an exponential avalanche of undetected cases. They will produce an exponentially growing number of detectable and medically serious cases. The contact tracing of those (upward in the infection tree) is tedious, and cannot fully eliminate the danger of such an avalanche.

Contact tracing as a main strategy thus only becomes viable once the value of *f*_asym_ is well established, and one is certain to be able to control the value of *R* such that *R f*_asym_ < 1.

## Appendix B Algorithm to simulate mitigation of reboot

### 1. Definitions

- *t* = 1, 2, … : Time in days (integer).
- *n*_int_: Number of interventions (including the reboot at *t* = 1).
- *t*_int_(*j*): First day on which the *j*’th rate *k*_*j*_ applies. On day *t*_int_(1) *≡* 1 the initial reboot step is taken.
- Δ*t*(*j*) = *t*_int_(*j* + 1) − *t*_int_(*j*): Time span between interventions *j* and *j* + 1.
- *t*_first_: First day on which the current rate *k* = *k*(*t*) is applied.
- *i*(*t*): Fraction of infected people on day *t*.
- *k*(*t*): Growth rate on day *t*.
- *r*: Number of tests per day.
- *C*_H_: Health cost.
- *C*_E_: Economic cost.
- *k*_min_= 0.005: Minimal growth rate targeted.
- *i*_low_ = 0.2: Lower threshold for *i/i*^*\**^. If *i/i*^*\**^ < *i*_low_, a relaxing intervention is made, irrespective of the estimate of *k*.
- *i*_high_ = 3: Upper threshold for *i/i*^*\**^. If *i/i*^*\**^ > *i*_high_, an intervention is made even if *k* is still smaller than *α δk*.
- *k*_low_ = 0.1: Minimal possible decreasing rate considered.
- *K*_high_ = 0.23: Maximal possible increasing rate considered.
- *T*_min_ = 3: Minimal time to wait since the last intervention, for interventions based on the level of *i*(*t*).
- *b*: Parameter defining the possible range of changes Δ*k* due to measures taken after estimating *k*. |Δ*k/k*_est_| ∈ [*b*, 1*/b*]. Usually we set *b* = 0.5.
- *α*: Confidence parameter.
- *N* (*t*): Cardinality of random sample of infected people on day *t*. The number *N* (*t*) is obtained by sampling from a Gaussian distribution of mean *i*(*t*) *r* and standard deviation 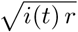 and rounding the obtained real number to the next non-negative integer.

### 2. Initialization

- *t*_first_ = *t*_int_(1) = 1.
- *n*_int_ = 1.
- *C*_H_ = 1.
- *C*_E_ = 0.
- *k*(1) = *k*_1_ = 0.1. (Initial growth rate)
- *i*(1) = *i*^*\**^. Common choice *i*^*\**^ = *i*_c_*/*4 = 0.0007.
- Draw *N* (1).
- *k*(2) = *k*(1). (No intervention at the end of day 1)
- Set *t* = 2.

### 3. Daily routine for day *t*

Define *i*(*t*) = *i*(*t* − 1) *e*^*k* (*t*−1)^,

Define *C*_H_ = max*{C*_H_, *i*(*t*)*/i*^*\**^*}*,

Define *C*_E_ = *C*_E_ *k*(*t*).

Draw *N* (*t*).

Determine what will be *k*(*t* + 1), by assessing whether or not to intervene:

**If** *t* = *t*_first_, **then** *k*(*t* + 1) = *k*(*t*). (No intervention)

**Else** Distinguish three intervention cases:

1. **If** *i*(*t*)*/i*^*\**^ < *i*_low_ and *t* − *t*_first_ ≥ *T*_min_, **then** *k*(*t* + 1) = min*{k*(*t*) + *x k*_1_, *k*_high_*}* with *x* = Unif[0, 1].
2. **ElseIf** *i*(*t*)*/i*^*\**^ > *i*_high_ and *t* − *t*_first_ ≥ *T*_min_, **then** *k*(*t* + 1) = max*{k*(*t*) − (1 + *x*)*/*2 *k*_high_, *k*_low_*}* with *x* = Unif[0, 1].
3. **ElseIf** *i*_low_ < *i*(*t*)*/i*^*\**^ < *i*_high_, **the**
  - set Δ*t ≡ t* − *t*_first_ + 1
  - Compute *k*_est_(*t*_first_, Δ*t*), and *δk*_est_(*t*_first_, Δ*t*) using Sec. B 4. **If** |*k*_est_| > *k*_min_ **AND** [*k*_est_ > *α δk*_est_ **OR** *k*_est_ < − *α δk*_est_], set *k*(*t* + 1) = *k*(*t*) − *x k*_est_ with *x* = Unif[*b*, 1*/b*]. If *k*(*t* + 1) > *k*_high_, put *k*(*t* + 1) = *k*_high_. If *k*(*t* + 1) < *k*_low_, put *k*(*t* + 1) = *k*_low_.
4. **Else** *k*(*t* + 1) = *k*(*t*)

*t* = *t* + 1.

**If** an intervention was taken above:

- Put *n*_int_ = *n*_int_ + 1.
- Define *t*_int_(*n*_int_) = *t* + 1.
- Define Δ*t*(*n*_int_ − 1) = *t*_int_(*n*_int_) − *t*_int_(*n*_int_ − 1).
- Set *t*_first_ = *t* + 1.

**If** |*k*_est_| < *k*_min_ **AND** *k*(*t*) < *k*_min_ **AND** *t* − *t*_first_ > 10, **then** EXIT.

**Else** Return to daily routine for next day.

### 4. Estimate of *k*(*t*, Δ*t*)

Computing *k*_est_(*t*_first_, Δ*t*) and *δk*_est_(*t*_first_, Δ*t*):

**If** Δ*t* is even:

Define

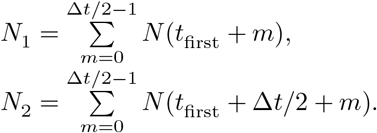

- **If** *N*_1_ *N*_2_ > 0, **then**

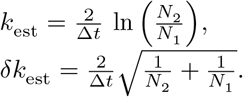
- **Else** return *k*_est_ = 0, *δk*_est_ = 1000.

**If** Δ*t* is odd:

Define

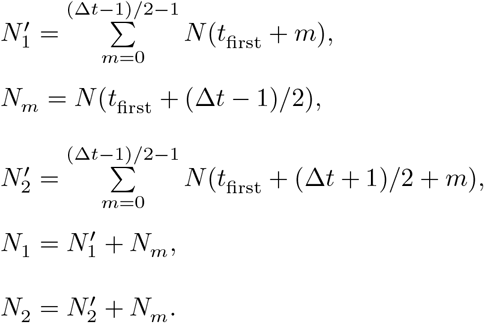

- **If** *N*_1_ *N*_2_ > 0, **then**

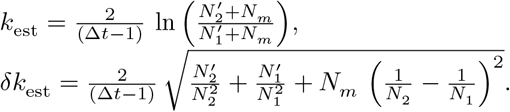
- **Else** return *k*_est_ = 0, *δk*_est_ = 1000.

### 5. Observables

Time to first intervention: Δ*t*(1) Health cost: *C*_H_

Political cost: *n*_int_

Economic cost *C*_E_

## Footnotes

1 As of early April 2020, according to the liveticker of the Swiss radio and television https://www.srf.ch/.

2 We prefer the terminology *physical distancing* to *social distancing*.

3 If the fraction of infected people can be measured via sewage water, *r* will be related to the number of people connected to a given sewage plant. But at this point, the relationship between such data and the actual current number of infections remains a topic for research. Of course, once key parameters such as the lag time between infection and incidence of biomarkers in sewage are known, sewage tests could become highly competitive.

4 Note that if tests take the nonvanishing time *t*test to yield a diagnosis, this time needs to be subtracted from the denominator in Eq. (2a), thereby resulting in an increase of the full testing rate 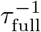.

5 It is important that the set of randomly selected people must change constantly, so that it should happen extremely rarely that a given person is tested twice.

6 Here, we solely focus on a person being infectious, but *not* on whether the person has developed antibodies. The latter test indicates that the person has been infected any time in the past. Serological tests for antibodies and (potential) immunity have their own virtue, but aim at different goals from the random testing for infections that we advocate here. By following the fraction of infections as a function of time, we can determine nearly instantaneously the growth rate of infections, *k*(*t*), and thus assess *and quantify* the effectiveness of socio-economic restrictions through the observed changes in *k* following a change in policy. This monitoring can even be carried out in a regionally resolved way, such that subsequently, restrictive or relaxing measures can be adapted to different regions (urban/rural, regions with different degrees of immunization, etc.).

7 A suppression of the COVID-19 pandemic is achieved if, for a sufficiently long time, the number of infections decays exponentially with time. Mitigation aims to reduce the exponential rate of growth in the number of infections. Stability is achieved when that number tends to a constant. Once stability is reached, one may start relaxing the restrictions step by step and monitor the effect on the growth rate *k* as a function of geographic regions.

8 Replacing the function *k*(*t*), assumed to be differentiable, by a piecewise constant function is a good approximation provided 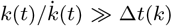 where 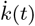 is the time derivative of *k*(*t*) and Δ*t*(*k*) is given by Eq. (13a) with the replacement *k*_1_ → *k*(*t*).

9 More precisely, 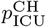 is the expected time (in Switzerland) for an infected person to spend in an intensive care unit (ICU) divided by the expected time to be sick.

10 If the infected fraction of people is *i*(*t*), its growth rate is 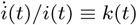 with the time derivative of *i*(*t*) denoted by 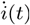.

11 One uses Eq. (7) to reach this conclusion.

12 One might also consider other distinguishing characteristics of groups (age or commuting habits, etc.), but we will not do so here, since it is not clear whether the increased complexity of the model can be exploited to reach an improved data analysis. In fact we expect that the number of fitting parameters will very quickly become too large by making such further distinctions.

